# Neurophysiological sensitivity in early childhood: The aperiodic EEG slope moderates the association between maternal anxiety and child internalizing symptoms

**DOI:** 10.1101/2025.11.01.25339315

**Authors:** Dashiell D. Sacks, Viviane Valdes, April R. Levin, Charles A. Nelson, Michelle Bosquet Enlow

## Abstract

Maternal internalizing (anxiety and depressive) symptoms are a robust risk factor for the development of internalizing symptoms in offspring, yet the neurobiological mechanisms that influence this association remain relatively unexplored. The aperiodic ‘slope’ of the EEG power spectrum (i.e., aperiodic exponent) is hypothesized to index the cortical excitatory-inhibitory balance and may serve as an early neurophysiological marker of mental health risk. In a prospective longitudinal cohort (*N* = 322 mother-child dyads), we examined associations among maternal anxiety and depressive symptoms in infancy and at age 5 years, child EEG aperiodic slope at age 3 years, and child internalizing symptoms at age 5 years. We investigated whether the aperiodic slope at 3 years (a) mediated associations between maternal internalizing symptoms in infancy and child internalizing symptoms at age 5 years and/or (b) moderated associations between maternal internalizing symptoms and child internalizing symptoms at age 5 years. There were no significant mediation effects. The aperiodic slope moderated the association between maternal anxiety symptoms and child internalizing symptoms: A steeper slope was associated with a stronger association between maternal and child symptoms. Findings suggest that the EEG aperiodic slope may represent a moderator of intergenerational risk for internalizing symptoms in early childhood.

## Introduction

Internalizing disorders, in particular anxiety, are among the most prevalent disorders in children, and can emerge as early as toddlerhood (Bufferd et al., 2011; Egger & Angold, 2006). Prevalence estimates in US youth range from 10% to 20% (Bufferd et al., 2011; Egger & Angold, 2006; Ghandour et al., 2019; Lu, 2019), although these estimates are likely conservative, as incidence and cases have steadily increased over time (Wu et al., 2023), with accelerated increases following the coronavirus disease 2019 (COVID-19) pandemic (Racine et al., 2021; Wu et al., 2023). Early onset of internalizing symptoms in childhood is associated with long-term chronicity, heterotypic continuity, and adverse socioemotional and economic outcomes (Beesdo et al., 2009; Vergunst et al., 2023). Extensive research indicates that children of caregivers with elevated internalizing symptoms are at elevated risk of developing internalizing psychopathology themselves (Goodman et al., 2011; Hentges et al., 2020). Identifying neurobiological mechanisms that influence the association between parent and child mental health may inform early identification and intervention strategies and improve long-term outcomes for children.

Guided by the Adaptive Calibration Model (ACM), we have recently investigated the role of child parasympathetic reactivity to a fearful stimulus, neural responses to emotional faces, and frontal alpha asymmetry at baseline in the intergenerational transmission of internalizing psychopathology (Kane-Grade et al., 2024; Quigley et al., 2023; Sacks, 2025). The ACM is an evolutionary-developmental model that provides an integrative framework for evaluating how children may calibrate biological stress-response systems in response to stress exposures. Within this framework, these calibrations may increase vulnerability through biological embedding of early adversity (*i.e.*, mediation). Such adaptations, in turn, may shape future responsivity to stress exposures (*i.e.*, moderation). The ACM can be applied to studies of the intergenerational transmission of internalizing symptoms, conceptualizing child exposure to maternal symptoms as a form of stress. Guided by this framework, Quigley et al. (2023) and Kane-Grade et al. (2024) tested mediation and moderation models, finding evidence that, at 3 years of age, heightened parasympathetic reactivity to a fearful video and neural responses to emotional faces, respectively, moderated the association between maternal and child internalizing symptoms at 5 years of age. Sacks (2025) found a mediation (but not moderation) effect, with baseline frontal alpha asymmetry at 5 years of age mediating the association between maternal symptoms at 3 years and child internalizing symptoms at 7 years. Together, these studies provide emerging evidence for the role of different physiological and neural processes in the intergenerational transmission of internalizing symptoms.

The aperiodic slope of the EEG power spectrum has recently received attention as a potential index of the balance between cortical excitation and inhibition (E-I balance) (Gao et al., 2017). Unlike traditional EEG power measures, which typically focus on rhythmic, oscillatory EEG activity and often conflate periodic and aperiodic components of the power spectrum, the aperiodic slope is derived from aperiodic EEG power (Donoghue et al., 2020). This aperiodic component follows a 1/f^x^-like distribution, in which power decreases as frequency increases. The exponent x (often represented as a linear slope in the log-log space) quantifies the rate of decrease; a smaller exponent (flatter slope) is hypothesized to reflect increased excitatory activity over inhibitory activity, and a larger exponent (steeper slope) to reflect increased inhibitory activity over excitatory activity. In our recent study, we found that in infancy, the aperiodic slope moderated associations between maternal internalizing symptoms (i.e., anxiety and depressive symptoms) and infant orienting/regulation, a core early temperament trait linked to later psychopathology (Sacks, Levin, et al., 2025). Further, in a separate study, we found a direct association between greater maternal anxiety and a flatter aperiodic slope at 3 years of age (Sacks, Valdes, et al., 2025). These findings suggest that aperiodic slope could reflect a neurobiological marker involved in the intergenerational transmission of internalizing symptoms.

Building on this emerging literature, in the current study we investigated the potential role of the aperiodic slope in the association between maternal and child internalizing symptoms in early childhood. We adopted a theoretical framework (Figure 1) building on our recent studies, Quigley et al. (2023), Kane-Grade et al. (2024) and Sacks et al., (2025), all of which were informed by the ACM. In line with this framework, the aperiodic slope may index a neurological adaptation to early exposure to maternal internalizing symptoms (*i.e.*, shift in E-I balance) that increases vulnerability to the development of subsequent internalizing symptoms. Furthermore, particularly in light of the moderating effects found in Sacks, Levin, et al. (2025), the aperiodic slope may reflect a neurobiological characteristic that influences sensitivity to maternal internalizing symptom exposure. No prior study has investigated whether the slope mediates or moderates the association between maternal and child internalizing symptoms. Accordingly, we aimed to investigate (1) whether child aperiodic slope at age 3 years mediated the association between maternal anxiety and depressive symptoms in infancy and child internalizing symptoms at age 5 years, and (2) whether child aperiodic slope at age 3 years moderated associations of maternal anxiety and depressive symptoms at age 5 years with child internalizing symptoms at age 5 years.

**Figure 1.**
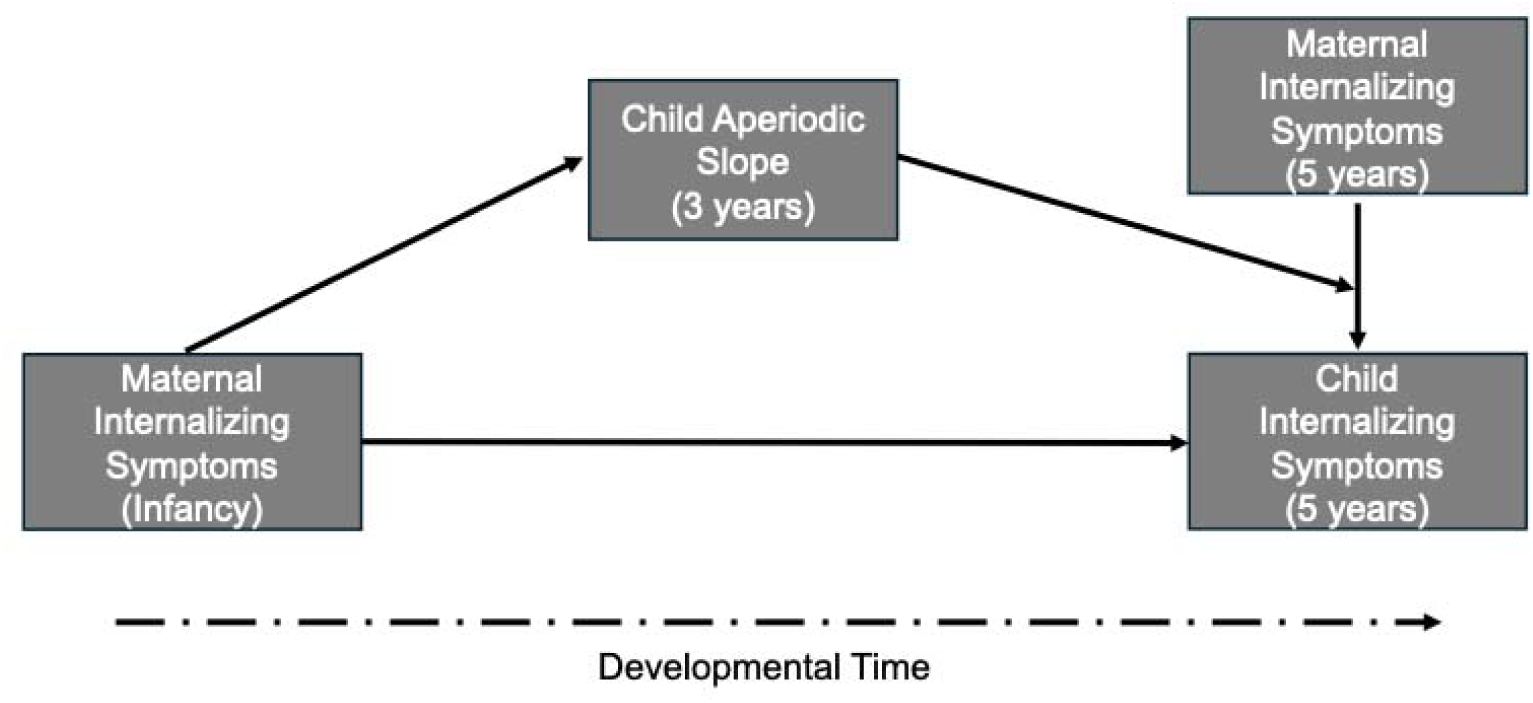
Conceptual model depicting the aperiodic slope (exponent) as a mediator and a moderator of the association between maternal and child internalizing symptoms.

## Methods

### Participants

Participants were recruited from a registry of local births comprising families that had indicated a willingness to participate in developmental research. Families in the current analyses participated in a prospective longitudinal study to examine the early development of emotion processing. Exclusion criteria included known prenatal or perinatal complications, maternal use of medications during pregnancy that may have a significant impact on fetal brain development (i.e. anticonvulsants, antipsychotics, opioids), pre- or post-term birth (±3 weeks from due date), developmental delay, uncorrected vision difficulties, and neurological disorder or neurological trauma. After enrollment, families were no longer followed, and their data were excluded from analyses if the child was diagnosed with an autism spectrum disorder or a genetic or other condition known to influence neurodevelopment. By design, families were enrolled in the parent study and completed initial assessments when the children were 5, 7, or 12 months of age (infancy), with a smaller subsample to be followed longitudinally for in-person assessments at 3 years and 5 years of age. Families were included in the current analyses if they had maternal internalizing symptom data at child age infancy and/or 5 years (*N* = 313), child baseline EEG data at 3 years (*N* = 168 retained after preprocessing), and/or child internalizing symptom data at age 5 years (*N* = 226), resulting in a total analytic sample of N = 322 mother-child dyads.

### Procedures

Mothers were asked to complete questionnaires via an online survey prior to or during laboratory visits. Questionnaires relevant to the current analyses included assessments of sociodemographic characteristics collected at enrollment in infancy, maternal anxiety and depressive symptoms (infancy and 5 years), and child internalizing symptoms (5 years). EEG data were collected during the 3-year laboratory visit. The Institutional Review Board at Boston Children’s Hospital approved all methods and procedures used in this study. Parents provided written informed consent prior to the initiation of study activities.

## Measures

### Sociodemographic characteristics

Sociodemographic characteristics were collected via parent report. Data collected included child age at each visit, sex assigned at birth (hereafter ‘sex’), race, and ethnicity, parent education, and annual household income (Table 1).

**Table 1.** Sample sociodemographic characteristics, collected at infancy (N=322).

| Variables | *n* | % |
| --- | --- | --- |
| Child sex |  |  |
| Male | 173 | 53.6 |
| Female | 150 | 46.4 |
| Child race |  |  |
| White | 260 | 80.5 |
| Black/African American | 7 | 2.2 |
| Asian American | 10 | 3.1 |
| More than one race | 42 | 13 |
| Not reported | 4 | 1.2 |
| Child ethnicity |  |  |
| Non-Latino/a, Spanish, or Hispanic | 284 | 87.9 |
| Latino/a, Spanish, or Hispanic | 35 | 10.8 |
| Not reported | 4 | 1.2 |
| Maternal educational attainment |  |  |
| Less than high school | 2 | 0.6 |
| High school/GED | 12 | 4.4 |
| Associate degree | 4 | 3.7 |
| Bachelor's degree | 93 | 28.8 |
| Master's degree | 147 | 45.5 |
| M.D., Ph.D., J.D., or equivalent | 63 | 19.5 |
| Not reported | 2 | 0.6 |
| Paternal educational attainment |  |  |
| Less than high school | 0 | 0 |
| High school/GED | 27 | 8.4 |
| Associate degree | 15 | 4.6 |
| Bachelor's degree | 95 | 29.4 |
| Master's degree | 102 | 31.6 |
| M.D., Ph.D., J.D., or equivalent | 79 | 24.5 |
| Not reported | 5 | 1.5 |
| Annual family household income |  |  |
| Less than \$35,000 | 8 | 2.7 |
| \$35,000-\$49,000 | 12 | 3.7 |
| \$50,000-\$74,999 | 32 | 9.9 |
| \$75,000-\$99,000 | 50 | 15.5 |
| \$100,000 or more | 199 | 61.6 |
| Not reported | 22 | 6.8 |

### Maternal anxiety symptoms (infancy, 5**D**years; predictor)

Maternal anxiety symptoms were measured at infancy and age 5 years via the Trait Anxiety form of the Spielberger State-Trait Anxiety Inventory (STAI-T; (Spielberger et al., 1983)). The STAI-T is a 20-item self-report questionnaire designed to measure anxiety proneness. Respondents are asked to rate the frequency of general mood states on a 4-point scale, ranging from ‘almost never’ to ‘almost always.’ Item scores were summed to create a total score (possible range: 20-80), with a higher score indicating greater trait anxiety. The STAI-T has established good internal consistency (α = .90), and test-retest coefficients from .73 to .86 (Barnes et al., 2002). Cronbach’s alpha scores in this sample were .89 and .91 at infancy and 5 years, respectively.

### Maternal depressive symptoms (infancy, 5**D**years; predictor)

Maternal depressive symptoms were measured at infancy and age 5 years via the Beck Depression Inventory (BDI-IA; (Beck et al., 1961). The BDI-IA is a 21-item self-report questionnaire that assesses the frequency and intensity of depressive symptoms in the past week. Items are scored on a 4-point scale (range 0-3) and summed to create a total score, with a total possible range of 0-63; higher scores indicate greater depressive symptoms. Internal consistency for the BDI has been reported as ranging from .73 to .92, with a mean of .86 (Beck et al., 1988). Cronbach’s alpha scores in this sample were .78 and .84 at infancy and 5 years, respectively.

### Child internalizing symptoms (5 years; outcome)

Mothers completed the Child Behavior Checklist 1.5-5 (CBCL/1.5-5) at the 5-year assessment (Achenbach & Rescorla, 2001). The CBCL forms are well-established, empirically supported questionnaires for assessing child psychopathology symptoms, producing scores on multiple syndrome and DSM-oriented scales, as well as higher-order symptom scores. The 99-item CBCL/1.5-5 asks parents to report on their children’s behavior during the past 6 months, with possible item scores ranging from 0 (“not true”) to 2 (“very true or often true”). The current analyses focused on the Internalizing Problems scale, comprising the following syndrome scales: Emotionally Reactive, Anxious/Depressed, Somatic Complaints, and Withdrawn. Cronbach’s alpha was .82 in this sample. Subscale raw scores are calibrated and normed by child age and gender, with normed scores expressed as the standard T-score metric (*M* = 50, *SD* = 10). A T-score < 60 is suggestive of a nonclinical level of symptoms, 60-63 of borderline clinical significance, ≥ 64 of clinical significance.

### EEG

#### EEG Acquisition

Continuous scalp EEG was recorded at 3 years using a 128-electrode HydroCel Geodesic Sensor Net (Electrical Geodesic Inc.). The net was connected to a NetAmps 300 amplifier (Electrical Geodesic Inc.) and referenced online to a single vertex electrode (Cz). Data were sampled at 500 Hz. Electrode impedances were kept at or below 100 kΩ, which is within recommended guidelines given the high-input impedance capabilities of the system’s amplifier. Participants sat on their caregiver’s lap in a dim room and watched a computer-generated video of moving infant toys while baseline EEG (2 minutes) was recorded.

#### Preprocessing

Raw Netstation (Electrical Geodesics, Inc) EEG files were exported into the MATLAB MAT-file format for preprocessing in MATLAB (version R2023a), using the Batch Automated Processing Platform (BEAPP; (Levin et al., 2018)), with integrated Harvard Automated Preprocessing Pipeline for EEG (HAPPE; (Gabard-Durnam et al., 2018)). Data were high-pass filtered at a 1Hz and low-pass filtered at 100Hz. Data were resampled to 250Hz, and then preprocessed using the HAPPE 1.0 module, which includes line noise removal using CleanLine multi-taper regression, bad channel rejection through evaluation of the normed joint probability of the average log power, and artifact removal using combined wavelet-enhanced independent component analysis (ICA) and Multiple Artifact Rejection Algorithm (MARA; (Winkler et al., 2015)). In addition to the 10-20 channels, the following channels were used for MARA: 28, 19, 4, 117, 13, 112, 41, 47, 37, 55, 87, 103, 98, 65, 67, 77, 90, 75. After artifact removal, bad channels were interpolated using spherical interpolation (with Legendre polynomials up to the 7th order). Data were referenced to the average reference, detrended to the signal mean, and segmented into 2-second epochs. HAPPE’s amplitude and joint probability criteria were used to reject epochs contaminated with artifact. Recordings with fewer than 20 segments (40 seconds of total EEG), percent good channels < 80%, percent independent components rejected >80%, mean artifact probability of components kept > 0.3, and/or percent variance retained < 25% were rejected (*N* = 5, 3%).

#### Power spectral density

Power spectral density was calculated in the BEAPP Power Spectral Density (PSD) module using a multitaper spectral analysis with three orthogonal tapers. For each channel, the PSD was averaged across segments, and then further averaged across all channels (10-20 and MARA; ‘33’, ‘22’, ‘9’, ‘122’, ‘28’, ‘24’, ‘19’, ‘11’, ‘4’, ‘124’, ‘117’, ‘13’, ‘112’, ‘45’, ‘41’, ‘36’, ‘37’, ‘55’, ‘87’, ‘104’, ‘103’, ‘108’, ‘47’, ‘52’, ‘67’, ‘62’,’77’, ‘92’, ‘98’, ‘58’, ‘65’, ‘70’, ‘75’, ‘83’, ‘90’, ‘96’).

#### Spectral Parametrization

The PSD was parameterized using the modified version of SpecParam (Donoghue et al., 2020) first reported by Wilkinson et al. (2024). This version was modified to improve model fit in early childhood. Specifically, in this version, the robust_ap_fit function is modified, so that the initial estimate of the flattened power spectra (flatspec) has a baseline elevated such that the lowest point is ≥ 0. Further details are available in Donoghue et al. (2020) and Wilkinson et al. (2024). The SpecParam model was used across a 2-55 Hz frequency range, in the fixed mode (no spectral knee), with peak_width_limits set to [0.5, 18.0], max_n_peaks = 7, and peak_threshold = 2. Mean R^2^ for the full sample was 0.997 (*SD* = 0.005). Mean estimated error for the full sample was 0.018 (*SD =* 0.011). The aperiodic exponent value was extracted for the current analyses. The aperiodic exponent is a positive value that reflects the rate of decline of the aperiodic slope, i.e., the exponent x in the 1/f^x^ distribution. A larger exponent value indicates a steeper slope, whereas a smaller exponent value indicates a flatter slope.

### Statistical Analysis

Statistical analyses were conducted in R 4.3.2. Descriptive statistics for the sample sociodemographic characteristics and main study variables were calculated. Pearson correlations among each of the main study variables were reported. Sex differences were tested among the main study variables, given established sex differences in internalizing and the aperiodic exponent (slope). Sex was included in subsequent analyses if significant differences among the main variables were established. Mediation and moderation analyses were conducted using the PROCESS Macro (André, 2021). Separate mediation models were investigated with maternal anxiety or depressive symptoms during infancy as the predictor, slope at 3 years as the mediator, and child internalizing symptoms at 5 years as the outcome. Individual paths and direct, indirect, and total effects were examined for each model. Significance of indirect effects was determined using bootstrapped (*n* = 5000) confidence intervals (95% CI). Additionally, two moderation models examined the main effects of slope at 3 years, maternal symptoms at 5 years, and the interaction effect of maternal symptoms and child slope on child internalizing symptoms at 5 years; maternal anxiety symptoms and depression symptoms were analyzed in separate models. Significant interaction effects were probed with conditional effects and floodlight analyses.

MissForest was used to impute missing data at each timepoint, for each variable in the mediation and moderation analyses (Stekhoven & Bühlmann, 2011). MissForest is a nonparametric, iterative imputation method based on Random Forests. MissForest initializes by fitting the data with the mean or mode of each variable and then iteratively averages over multiple unpruned classification or regression trees until arriving at a solution. Supplementary analyses were conducted for model comparison in the subset of participants with complete data for all the model variables (*N* = 113 for mediation, *N* = 120 for moderation).

## Results

Sample sociodemographic characteristics are presented in Table 1. Descriptive statistics for the main study variables are presented in Table 2. Bivariate associations among the main study variables and sex are presented in Table 3. There were significant associations among each of the maternal and child psychopathology symptom variables. The aperiodic exponent was not significantly associated with maternal anxiety or depressive symptoms at infancy or 5 years or with child internalizing symptoms at 5 years. The aperiodic exponent differed by child sex (larger exponent among females; *p* <.05) and thus sex was included as a covariate in each of the main models.

**Table 2.** Descriptive statistics for the main study variables.

|  | Min | Max | Mean | SD |
| --- | --- | --- | --- | --- |
| Maternal anxiety symptoms, Infancy | 20 | 54 | 33.36 | 7.13 |
| Maternal anxiety symptoms, 5 years | 20 | 58 | 32.94 | 7.86 |
| Maternal depressive symptoms, Infancy | 0 | 22 | 5.28 | 3.86 |
| Maternal depressive symptoms, 5 years | 0 | 24 | 4.88 | 4.34 |
| Aperiodic exponent, 3 years | 0.92 | 1.44 | 1.13 | 0.10 |
| Child internalizing T-score, 5 years | 29 | 72 | 46.43 | 9.55 |

**Table 3.** Bivariate associations among the main study variables.

|  | STAI – Inf | BDI – Inf | Exponent –<br>3Y | STAI – 5Y | BDI – 5Y | CBCL – 5<br>years |
| --- | --- | --- | --- | --- | --- | --- |
| BDI – Inf | .58*** | -- |  |  |  |  |
| Exponent – 3 years | -0.10 | -.11 | -- |  |  |  |
| STAI – 5 years | .70*** | .48*** | -.02 | -- |  |  |
| BDI – 5 years | .45*** | .53*** | .05 | .65*** | -- |  |
| CBCL – 5 years | .22** | .32*** | .07 | .26*** | .24*** | -- |
| Sex (Female) | -.03 | -.01 | .19* | .02 | .00 | .09 |

*Note.* STAI = Trait Anxiety form of the Spielberger State-Trait Anxiety Inventory (maternal anxiety symptoms; STAI-T); BDI = Revised Beck Depression Inventory (maternal depressive symptoms; BDI-IA); CBCL = Internalizing Problems T-score from the Child Behavior Checklist 6-18 years (child internalizing symptoms).
\* $p < .05$ , \*\* $p < .01$ , \*\*\* $p < .001$
Larger exponent indicates a steeper slope.

### Mediation Analyses

#### Maternal Anxiety Symptoms

A mediation analysis was conducted to examine whether child EEG slope at 3 years mediated the association between maternal anxiety symptoms in infancy and child internalizing symptoms at 5 years. There was no significant mediation effect. The indirect effect of maternal anxiety symptoms on child internalizing symptoms through EEG slope was not significant, 95% CI [-0.020, 0.013]. The direct effect of maternal anxiety symptoms on child internalizing symptoms remained significant (*p* < .001). See Table 4 for full mediation analysis results.

**Table 4.**
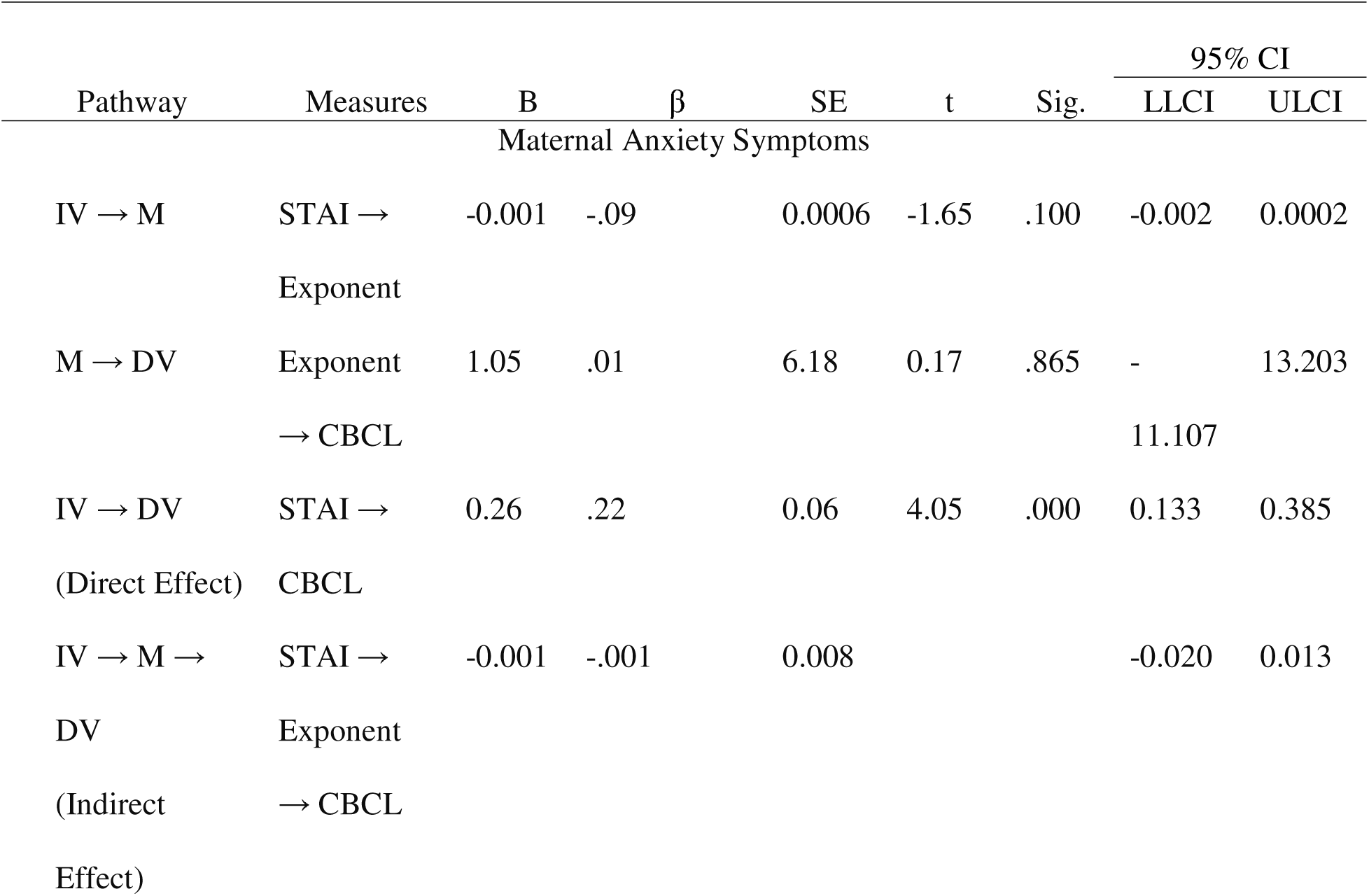
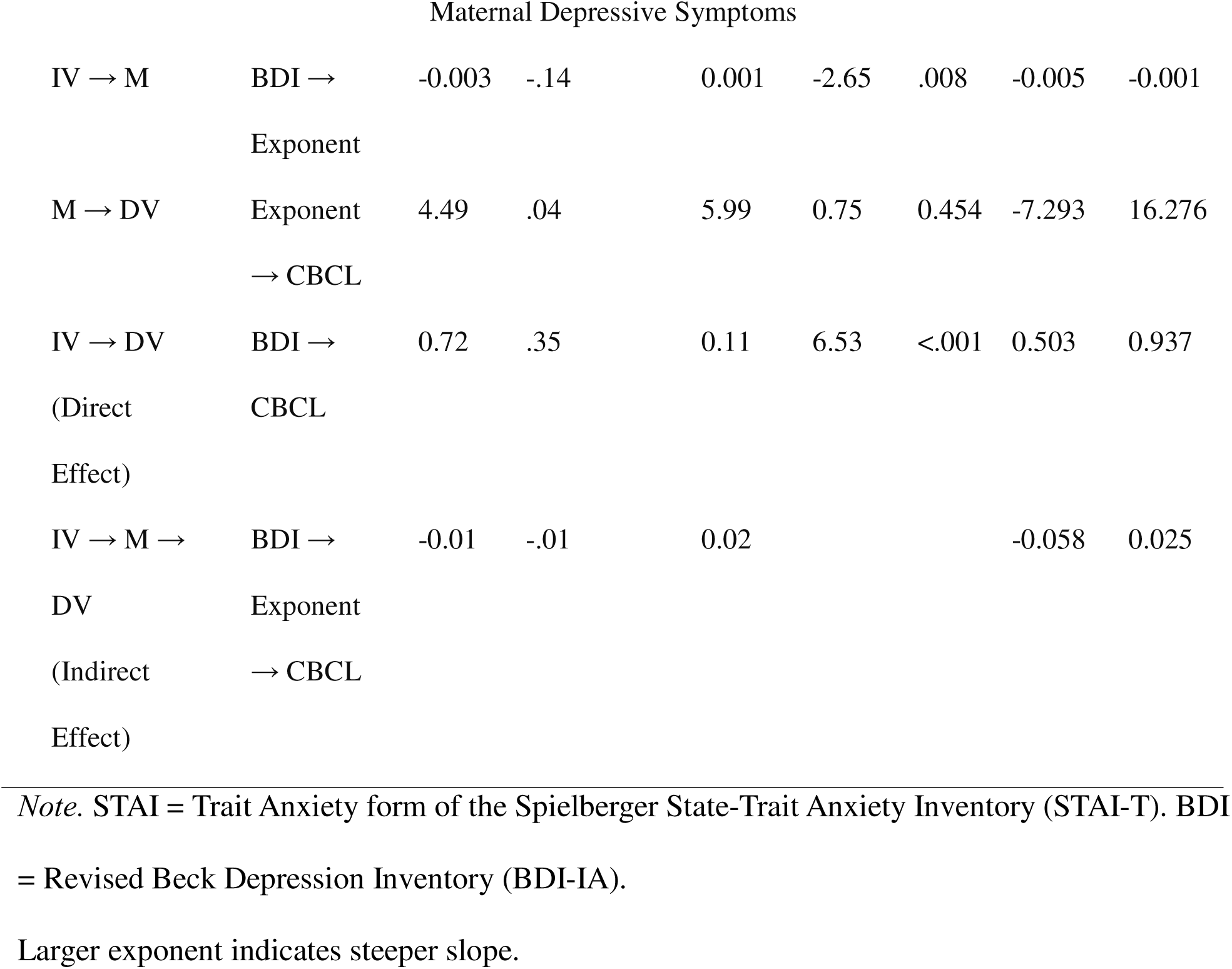
Mediation models for maternal anxiety symptoms and for depressive symptoms in infancy, child EEG slope at 3 years, and child internalizing symptoms at 5 years.

#### Maternal Depressive Symptoms

A separate mediation analysis was conducted to examine whether child EEG slope at 3 years mediated the association between maternal depressive symptoms in infancy and child internalizing symptoms at 5 years. There was no significant mediation effect. The indirect effect of maternal depressive symptoms on child internalizing symptoms through EEG slope was not significant, 95% CI [-0.058, 0.025]. The direct effect of maternal depressive symptoms on child internalizing symptoms remained significant (*p* < .001). See Table 4 for full mediation analysis results.

### Moderation Analyses

#### Maternal Anxiety Symptoms

A moderation analysis was conducted to test whether child EEG slope at 3 years moderated the association between maternal anxiety symptoms and child internalizing symptoms at 5 years, controlling for child sex (Table 5). The overall model was significant, R² = 0.10, F(4, 318) = 8.86, *p* < .001. There was a significant interaction effect (β = .20, *p* < .001), whereby the magnitude of the positive association between maternal anxiety symptoms and child internalizing symptoms was greater at higher values of the aperiodic slope (steeper slope). Conditional effects are presented in Table 6 and visualized in Figure 3, with the floodlight analysis.

**Table 5.** Regression model for child EEG exponent at 3 years and maternal anxiety symptoms at 5 years predicting child internalizing symptoms at 5 years.

| | B | $\beta$ | SE | t | p | LLCI | ULCI |
| --- | --- | --- | --- | --- | --- | --- | --- |
| Child EEG exponent | 0.345 | .003 | 6.01 | 0.06 | .954 | -11.479 | 12.169 |
| Maternal anxiety symptoms (STAI) | 0.267 | .23 | 0.065 | 4.077 | **<.001** | 0.138 | 0.395 |
| STAI x exponent | 3.10 | .20 | 1.11 | 2.79 | **.006** | 0.911 | 5.288 |
| Child sex | 1.14 | .14 | 0.91 | 1.25 | .212 | -0.653 | 2.934 |

*Note.* STAI = Trait Anxiety form of the Spielberger State-Trait Anxiety Inventory (STAI-T).
Larger exponent indicates steeper slope.

**Table 6.** Conditional effects for moderation by child EEG exponent (slope) at 3 years of the association between maternal anxiety symptoms and child internalizing symptoms at 5 years.

| Exponent | Effect | $\beta$ | SE | t | p | LLCI | ULCI |
| --- | --- | --- | --- | --- | --- | --- | --- |
| -0.075 | 0.03 | .03 | 0.12 | 0.27 | .785 | -0.205 | 0.271 |
| 0 | 0.27 | .23 | 0.07 | 4.08 | <.001 | 0.138 | 0.395 |
| 0.075 | 0.50 | .43 | 0.09 | 5.59 | <.001 | 0.324 | 0.676 |

*Note.* Larger exponent indicates steeper slope.

**Figure 3.**
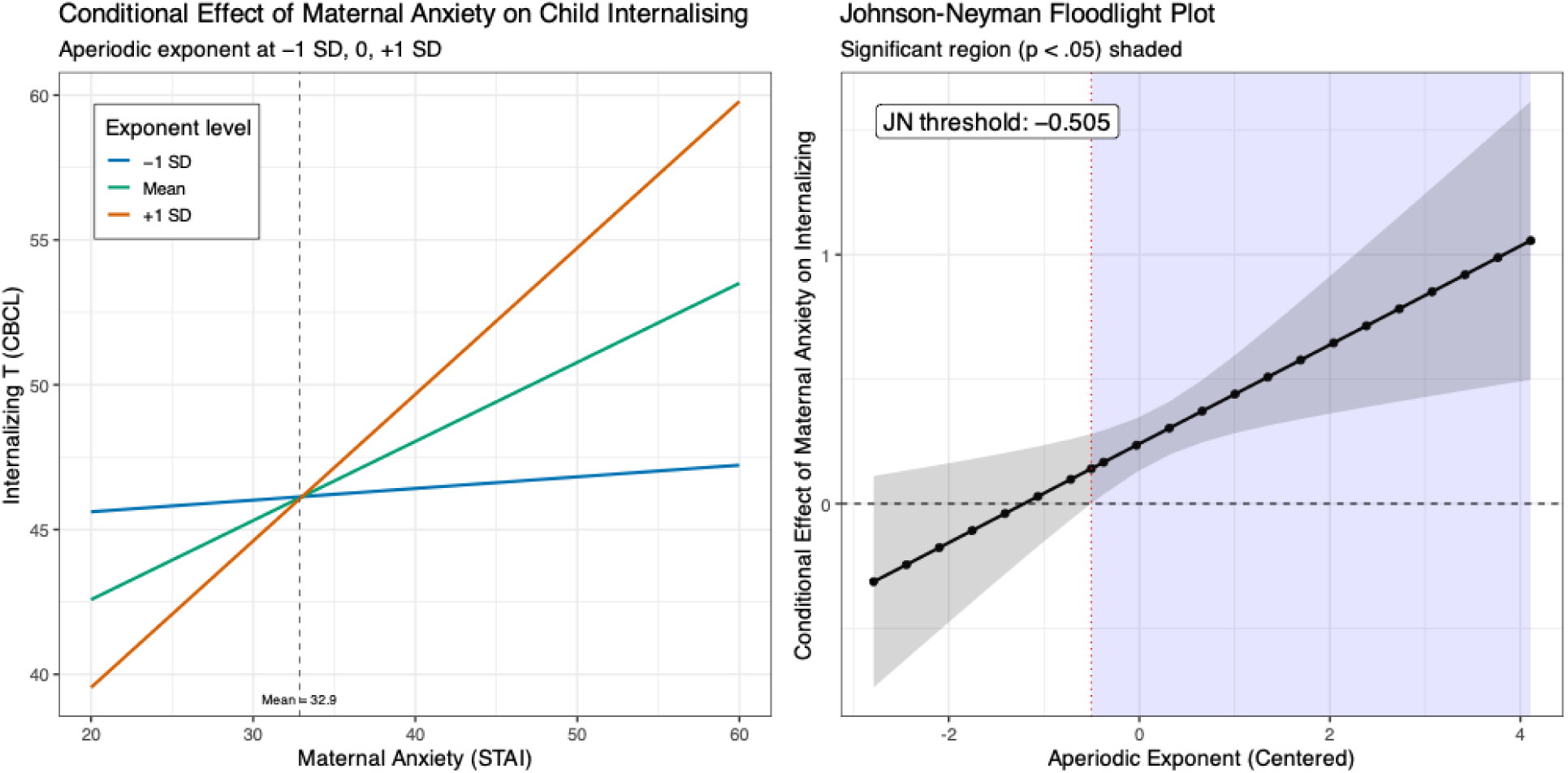
Moderating effect of the child aperiodic exponent (slope) at 3 years on the association between maternal anxiety symptoms and child internalizing symptoms at 5 years. Note. Conditional effects are presented on the left and Johnson-Neyman floodlight analysis on the right. To aid interpretation, the X-axis in the left panel is shown with raw STAI-T units (dashed line = sample mean).

#### Maternal Depressive Symptoms

A moderation analysis was conducted to test whether child EEG exponent (slope) at 3 years moderated the association between maternal depressive symptoms and child internalizing symptoms at 5 years (Table 7). The overall model was significant, R² = 0.092, F(4, 318) = 8.09, *p* < .001. The child EEG slope × maternal depressive symptoms interaction term was not significant, *p* = .241. Maternal depressive symptoms were independently associated with child internalizing symptoms, *p* < .001.

**Table 7.** Regression model for child EEG exponent (slope) at 3 years and maternal depressive symptoms at 5 years predicting child internalizing symptoms at 5 years.

| | B | $\beta$ | SE | t | p | LLCI | ULCI |
| --- | --- | --- | --- | --- | --- | --- | --- |
| Exponent | -0.78 | -.01 | 6.07 | -0.13 | .898 | -12.715 | 11.151 |
| Maternal depressive symptoms (BDI) | 0.56 | .27 | 0.11 | 4.91 | **<.001** | 0.335 | 0.783 |
| BDI x Exponent | 1.98 | .07 | 1.71 | 1.16 | .248 | -1.385 | 5.351 |
| Sex | 1.52 | .18 | 0.92 | 1.64 | .101 | -0.298 | 3.329 |

*Note.* BDI = Revised Beck Depression Inventory (BDI-IA).
Larger exponent indicates steeper slope.

#### Supplementary Analyses

Full model details for the supplementary analyses are provided in the Supplementary Materials. In summary, the results for all mediation and moderation models remained consistent in the subset of participants with complete data across time points (*i.e.*, no imputation).

## Discussion

In the present study, we investigated the potential mediating and moderating roles of the EEG aperiodic slope at 3 years on the intergenerational transmission of internalizing (anxiety and depressive) symptoms in a longitudinal sample. The aperiodic slope is a novel biomarker hypothesized to index the cortical E-I balance, with steeper slopes linked to greater inhibitory vs. excitatory activity, and flatter slopes linked to greater excitatory over inhibitory activity (Gao et al., 2017). There were no significant mediating effects: Associations between maternal symptoms at infancy and child slope at 3 years and between child slope at 3 years and child symptoms at 5 years were not significant. There was a significant moderating effect, with slope at 3 years moderating the association between maternal anxiety symptoms and child internalizing symptoms at 5 years.

The moderating effect observed in the present study builds on findings from our prior work in infancy (Sacks, Levin, et al., 2025), in which we found that the aperiodic slope moderated associations between maternal internalizing symptoms and infant orienting/regulation, with a steeper slope associated with a stronger negative association between maternal internalizing and infant orienting/regulation capacity. In the current study, we found evidence for a similar moderating effect, whereby a steeper aperiodic slope at 3 years was associated with a stronger association between greater maternal anxiety symptoms and greater child internalizing symptoms at 5 years. This finding aligns with our hypothesis that slope may index a neurobiological endophenotype of environmental sensitivity. This interpretation is consistent with the theory of differential susceptibility, which posits that individual variability in environmental sensitivity is driven by underlying neurobiological traits that emerge early in development (Boyce, 2016). Balanced E-I dynamics are thought to be necessary for maintaining cortical networks near criticality, which in turn supports maximum information transmission and capacity through the emergence of neural avalanches (Shew et al., 2011). Complementary theoretical and computational models suggest that E-I balances are essential for robust and selective neural responses in the presence of noise, supporting high-capacity, stable information processing. However, although emerging evidence supports aperiodic slope as a proxy for E-I balance, this remains a working theoretical framework. Thus, future research may refine or reinterpret the functional significance of slope, offering alternative explanations for the observed moderation effects that need to be considered.

The current results parallel the moderating effects observed our recent studies, Quigley et al. (2023) and Kane-Grade et al. (2024), in which we investigated parasympathetic reactivity to a fearful video and neural responsivity to emotional faces, respectively. These studies each investigated distinct neural and physiological processes within similar frameworks informed by the Adaptive Calibration Model (ACM). Taken together, the results across studies may reflect distinct, but converging pathways that influence variability in sensitivity to maternal psychopathology. Future studies may consider how these different systems interact to influence the intergenerational transmission of internalizing symptoms. In Quigley et al. (2023), parasympathetic reactivity moderated the association between both maternal depression and anxiety and child internalizing symptoms, whereas in Kane-Grade et al. (2024) and the present study, there was a significant moderating effect for maternal anxiety only. Despite comorbidity between maternal anxiety and depressive symptoms, they may have differential effects on parenting behavior (Quigley et al., 2023). The differences observed in parenting behaviors between parents with predominantly anxious versus depressive presentations may interact with child biological risk and protective factors distinctly, contributing to specificity in moderating effects. However, in our infancy study, we found a moderating effect of the aperiodic slope for the association of both maternal anxiety and depressive symptoms with infant orienting/regulation. Younger children may be more broadly sensitive to affective cues in the caregiving environment, or neurodevelopmental systems may be less differentiated early in development. Further research is required to investigate potential timing effects and other factors that may influence the associations. Given that this was a community sample with moderate symptoms levels, future research in clinical populations may also yield greater insight into differential effects across distinct psychopathology domains.

There were no significant mediating effects in the current study. This result may support the hypothesis that, early in development, the aperiodic slope reflects a relatively stable, trait-like characteristic, rather than a mechanism through which the environment influences the development of child psychopathology symptoms. However, the aperiodic slope has been shown to follow a nonlinear developmental trajectory across childhood (McSweeney et al., 2023; Sacks, Valdes, et al., 2025), with differential associations between maternal anxiety and slope at different ages and trajectories. More complex models may be required to fully capture interrelations among maternal input, child slope, and child psychopathology. Alternatively, given rapid development in early childhood, infancy to 5 years may reflect a relatively large developmental window for capturing more nuanced temporal associations. Study designs with repeated measurement within narrower intervals could yield more precise insights into potential associations among the aperiodic slope and maternal and child internalizing symptoms.

Strengths of this study include the longitudinal design with measures from infancy to 5 years of age. However, the findings should be considered in the context of its limitations. The sample comprised families who are predominantly White and of middle to high socioeconomic status, which may reduce the generalizability of findings. Levels of maternal and child psychopathology symptoms were moderate, and thus the nature of associations could vary in samples with clinical levels of internalizing symptoms. Both maternal and child internalizing symptom scales were maternal report, which could inflate associations or introduce bias, leading to inflated correlations. However, the results were moderated by EEG, an objectively recorded biological measure, and recent research suggests that mothers’ psychopathology minimally biases their ratings of their children’s emotions and behaviors (Olino et al., 2021). Thus, the moderation results are unlikely to be explained by issues related to maternal report. Finally, the aperiodic slope is a relatively novel measure, and best practices for calculation in early childhood are still being established. Here, we adopted the same processing parameters as recent studies (Sacks, Levin, et al., 2025; Sacks, Valdes, et al., 2025; Wilkinson et al., 2024). We focused on the slope averaged across the whole scalp. Future studies may consider more nuanced region-based analyses to investigate whether these associations vary by brain region, although focusing on whole scalp slope is consistent with most prior research in this area.

## Conclusion

The present study provides novel evidence for the aperiodic slope as a moderator of the intergenerational transmission of internalizing symptoms in early childhood. Guided by the ACM, we investigated both potential moderating and mediating effects, finding support for moderation. Specifically, the aperiodic slope at 3 years moderated the association between maternal anxiety and child internalizing symptoms at 5 years. The aperiodic slope has been proposed as a proxy measure for cortical E-I balance and, in line with the theory of differential susceptibility, may serve as a marker of environmental sensitivity, including to maternal caregiving behaviors in early childhood. Together with other biological markers identified in recent research, including parasympathetic reactivity and neural responses to emotional faces, the aperiodic slope may be a useful marker for identifying children with greater risk in early childhood and for developing targeted interventions aimed at reducing the risk for developing internalizing psychopathology.

## Data Availability

All data produced in the present study are available upon reasonable request to the authors

